# Efficacy and Safety of Cannabis Transdermal Patch for Alleviating Psoriasis Symptoms: Protocol for a Randomized Controlled Trial (CanPatch)

**DOI:** 10.1101/2023.03.08.23286926

**Authors:** Pim Sermsaksasithorn, Pravit Asawanonda, Phanupong Phutrakool, Thunnicha Ondee, Pajaree Chariyavilaskul, Sunchai Payungporn, Krit Pongpirul, Nattiya Hirankarn

## Abstract

**Background:** Existing topical psoriasis treatments are partially effective or have long-term side effects for a proportion of people with psoriasis; therefore, effective and safe treatment options are required. Cannabidiol, a cannabinoid in *Cannabis sativa*, reverses the etiology of psoriasis through skin receptors according to in vitro research. Cannabidiol transdermal patches may be an effective treatment for psoriasis, although the efficacy and safety data are limited.

**Methods and analysis:** This is a randomized double-blind controlled trial comparing cannabidiol (CBD) with minimal tetrahydrocannabinol (THC) patches with placebo patches (1:1 ratio) daily applied to comparable lesions of each patient with mild to moderate plaque-type psoriasis performed in a university hospital in Thailand (n=60). The primary outcome is the local psoriasis severity index (LPSI). The local severity index of psoriasis, the itch score using the visual analog scale, and adverse events will be evaluated on day 0, 30, 60, and 90 of the study. Furthermore, on days 0 and 90 of this study, biological samples will be taken for the evaluation of the skin, gut, and mouth microbial profile of 50% of randomly selected individuals.

**Conclusions:** This study aims to investigate the efficacy and safety of cannabidiol transdermal patches in alleviating the symptoms of psoriasis. We will also examine personal impacts on the efficacy and safety of patches, such as the microbial profile. The results of this study may highlight a novel topical treatment option that reduces suffering in patients with psoriasis.

**Ethics and dissemination:** This study was registered with the. The protocol is being considered by the Institutional Review Board of the Faculty of Medicine, Chulalongkorn University. The results of this study will be faithfully presented through conferences or published articles.

## INTRODUCTION

Psoriasis is a common chronic immune-mediated systemic disease with physical, psychological, and social burden due to its disfiguring effects and substantial comorbidities, including psoriatic arthritis, metabolic diseases, inflammatory bowel disease, cardiovascular diseases, and psychiatric disorders [1-4]. The prevalence of this disease ranges from 0.27% to 11.4% in various countries and tends to increase [5, 6].

The pathophysiology of psoriasis is multifactorial, but the essential part is immune-mediated inflammation with IL-17 and IL-23 as key players that result in uncontrolled proliferation and dysfunctional differentiation [1, 7]. Interestingly, recent studies have shown a significant difference between the human microbiota of patients with psoriasis and the normal population, with particular genera and species of skin microbiota clearly identified on the lesional skin and disproportions of the gut microbiota that influence the pathogenesis of psoriasis via the gut-skin axis [8-16].

Most patients have psoriasis vulgaris or plaque-type psoriasis with mild to moderate severity; therefore, topical therapies are the treatment of choice. However, in some individuals, conventional topical therapy options are ineffective or accompanied by adverse effects [7, 17-23]. For these reasons, novel topical therapies are warranted.

Emerging studies in recent decades have suggested that cannabidiol (CBD), a non-addictive cannabinoid of *Cannabis sativa*, reverses the primary pathogenesis of psoriasis through various skin receptors, including inhibiting keratinocyte proliferation, reducing inflammation, modifying Th1-Th2 balance, and inhibiting IL-17 [24-36]. However, limited clinical studies have been conducted.

### Cannabidiol (CBD) and psoriasis

Cannabinoids were found to exert opposite effects on the pathogenesis of psoriasis through skin receptors, including the CB1, CB2, GPCR55, TRPV1 and PPAR γ receptors [24-29, 37]. CBD, a cannabinoid derived from *Cannabis sativa* that is highly effective with minimal addictive properties, inhibits keratinocyte proliferation, reduces inflammation by inhibiting the function of NF-kB and TNF-α, modifies the balance of Th1-Th2, and alters cytokines by inhibiting IL-17 and IFN-γ [24, 27, 28, 30, 37-42].

By affecting CB1, CB2, and TRP channels in cutaneous nerve fibers, mast cells, and keratinocytes, cannabinoids also reduce itching. [38, 43, 44] In clinical trials, topical use of CBD reduced itching in patients with atopic dermatitis. [45, 46]

CBD is added to a variety of skin products, including those for psoriasis. [34, 38]. Despite a small number of clinical research studies, the current findings of the transdermal use of CBD ointment in the treatment of psoriasis are encouraging, including a significant reduction in the Psoriasis Area Severity Index (PASI) score on day 90 and the LPSI score on week 12 after the application of CBD ointment. [30, 34, 47]

### Cannabidiol (CBD) transdermal patch

Transdermal patches have been widely used in transdermal drug delivery systems. [48] The advantages of transdermal patches include non-invasiveness, painlessness, avoidance of first-pass metabolism, sustained medication release, and lack of gastrointestinal adverse effects [48-50]. Furthermore, transdermal patches provide better medication control and less dose variability than other transdermal drug delivery methods, such as ointments [48]. However, the lipophilic nature of cannabis makes formulation and absorption challenging for therapeutic impact, including poor skin diffusion [50-52]. Medium chain triglycerides (MCTs) have been found to be pharmaceutical carriers of lipophilic drugs [53] and used for cannabis preparation in both in vitro and in vivo studies [54-57].

### Safety of Cannabidiol (CBD)

Cannabidiol accumulates in organs of high blood supply such as the heart, brain, liver, and lungs when it enters the body [50]. The serious adverse events of CBD are hepatocellular damage, pneumonia, cardiovascular disease, and decreased fertility [58, 59]. However, many of the damage mentioned and alterations in vitro cell viability occurred at doses of more than 200 mg/kg/day [60]. Appetite loss, diarrhea, drowsiness, and sedation are other side effects [58, 59]. Chronic CBD causes the accumulation of CBD in adipose tissue, resulting in weeks of lethargy and sedation [50]. With the topical application of CBD, previous studies have not reported serious adverse events. Minorities experience minor adverse events, including erythema, skin irritation or pain, changes in hair or skin color, somnolence, and diarrhea [31, 32, 61-66].

### Gut, skin, and oral microbiota in psoriasis

Most of the research indicated significant changes in the microbial profile in patients with psoriasis. Substantial changes in overall biodiversity, decreased α-diversity, and increased β-diversity of the skin microbiota were discovered in psoriatic skin compared to healthy and non-lesional skin [8, 12, 67, 68]. The discovery of the hypothesis of the gut-skin axis stated that gut microbial dysbiosis leads to increased inflammatory cytokines and lessened tight junction integrity that allows blood and skin access to bacterial components and metabolite products [8, 11, 13, 15, 69, 70]. Subsequently, the studies revealed a reduction in the overall diversity of intestinal microbials and significant changes in β-diversity of the intestinal microbiome diversity in patients with psoriasis [15, 16, 68, 70, 71].

Some taxonomy of the microbiota distinguishes psoriasis from normal presentation. Compared to normal skin, the combined relative abundance of *Streptococcus spp*., *Staphylococcus spp*. and *Corynebacterium spp*. increases in psoriatic skin, while the abundance of *Staphylococcus epidermidis, Cutibacterium acnes*, and *Cutibacterium granulosum* decreases [11, 12, 72]. The Firmicutes to Bacteroidetes ratio (F/B ratio), two main phyla of the human gut microbiota, increases in patients with psoriasis, directly correlated with PASI scores and psoriasis comorbidities [8, 13-16]. Augmentation of *Ruminococcus gnavus* increased and underrepresentation of *Faecalibacterium prausnitzii* and *Akkermansia muciniphila* were also found in the intestinal microbiota of patients with psoriasis [8-10, 13, 15, 16, 70, 73].

For the oral microbiome, the interaction with psoriasis pathogenesis is not well established. Compared to healthy controls, patients with psoriasis are 1.55 times more likely to develop periodontal disease, a shared mechanism involving elevated Th-17 cells and IL-17 [74]. The microbiome was distinct in the saliva of periodontal disease [75-77]. No significance was found in α- or β-diversity of the oral microbiome of patients with psoriasis compared to normal people, although 18 species did [71].

## OBJECTIVES AND HYPOTHESES

Given the findings of a brief review of the literature showing promising therapeutic effects of CBD on psoriasis, we hypothesized that CBD with minimal transdermal patches of THC would be an effective and safe treatment option for psoriasis. Therefore, the main objective of this trial is to compare the efficacy of alleviating CBD psoriasis symptoms with minimal transdermal THC patches with placebo patches among patients with mild to moderate plaque-type psoriasis. Secondary objectives are to (1) report adverse events of CBD with minimal THC transdermal patches in patients with psoriasis (2) explore the microbial alteration of the gut, skin, and oral microbiome in patients with psoriasis who receive CBD with minimal THC transdermal patches and the relationship between the microbial profile and the clinical presentation of those patients.

## MATERIAL AND METHODS

### Study design

This is a randomized, double-blind, controlled study. This study is being approved by the Chulalongkorn University Faculty of Medicine Institutional Review Board (number XX) and registered with the Thai Clinical Trials Registry (TCTR No. 20220518004).

### Participants

Participants will be recruited through direct encounter and telephone contact. The participants will be from the Division of Dermatology of King Chulalongkorn Memorial Hospital’s database. Furthermore, the online poster will be advertised (e.g., on Facebook). Participants in this trial will be selected based on medical history, medical records, and physical examination by physicians to verify if they meet eligibility criteria and consent forms in accordance with the research ethics requirements outlined in the Declaration of Helsinki. Inclusion and exclusion criteria are shown in **Table 1**.

**Table 1.** Inclusion and exclusion criteria.

| Inclusion criteria | Exclusion criteria |
| --- | --- |
| - Aged 18 years and older<br>- Have been diagnosed with mild to moderate plaque-type psoriasis with a baseline PASI score of less than ten by a board-certified dermatologist<br>- Have two or more plaques of psoriasis of comparable size and severity<br>- Refrain from using topical psoriasis therapies for at least two weeks prior to research.<br>- Able and willing to give written informed consent | - Pregnant or breastfeeding<br>- Have contraindications to cannabis transdermal patches (Known allergy to medical cannabis, or patches' ingredients, history of severe liver or kidney disease, history of schizophrenia or other serious psychiatric diseases)<br>- Addictive behavior is defined as cannabis, opioids, or other recreational or pharmaceutical drug abuser (Determined by patient interview and medical records)<br>- Receive biologic therapy for psoriasis treatments<br>- Have had less than 12 weeks of conventional systemic therapy for psoriasis (oral medications or phototherapy)<br>- Have received conventional systemic therapy for psoriasis (oral medications or phototherapy) for more than 12 weeks but have seen variations in psoriasis symptoms or adjustments in their medication or phototherapy regimen within the past 12 weeks |

### Study medication

Intervention patches (P patches) will include 1.0 ± 0.1 mg of CBD in an MCT solvent on 2.0 ± 0.2 g of yellow adhesive layer covered with aluminum foil whereas placebo control patches (C patches) will be 2.0 g of yellow adhesive layer covered with aluminum foil.

### Procedures

All participants who meet the eligibility criteria and provide their written consent are invited to respond to the baseline demographic and clinical characteristics. For each participant, two similar psoriasis lesions are selected and then randomly assigned to the intervention plaque or the control plaque in a 1:1 ratio. An overview of the progress of the current trial is shown in the consolidated standards of reporting trials diagram (**Figure 1**). The study and control participants are instructed to apply a P or C patch, respectively, on a selected plaque of psoriasis every day for 90 days and at least 6 hours a day.

**Figure 1.**
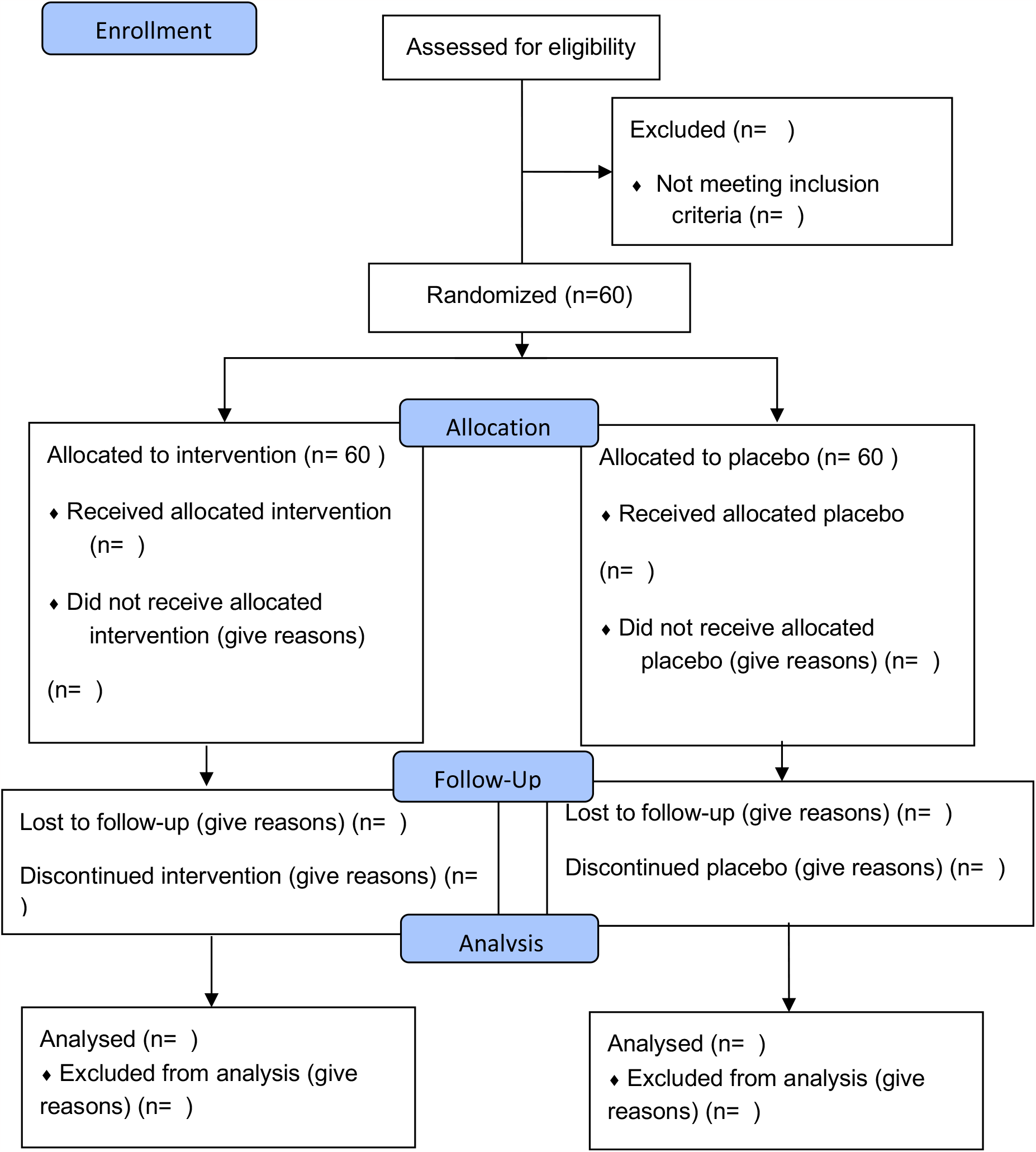
CONSORT flow diagram. flowchart of phases of Efficacy and Safety of Cannabis Transdermal Patch for Alleviating Psoriasis Symptoms: A Randomized Controlled Trial (CanPatch)

#### Adherence

Face-to-face reminder sessions are conducted at initial product dispensing and three subsequent visits. This session reviews the application of transdermal patches, including scheduling, storage, and missing dosage solutions. At each follow-up appointment, unused patches and used patches’ packages are tallied and documented.

#### Randomization

Two similar psoriasis lesions of each patient are randomly assigned to the intervention group or control group by block randomization with a ratio 1: 1 with blocks of 4 by STATA version 15.0 (StataCorp. 2017. Stata Statistical Software: Release 15. College Station, TX: StataCorp LLC.).

#### Blinding

After double-blind randomization, individuals not involved in the study translate the randomization code into instructions on which patches should be used on which lesion. They place the instructions in sealed envelopes that are then delivered to patients randomly by a physician. Each lesion is instructed in the same type of patch every day. The front of an opaque sealed envelope is the study ID and the date the envelope will be opened. Therefore, patients and physicians are blinded. Allocation concealment is ensured, as the service will not release the randomization code until data collection has been completed. A code might be broken in extraordinary cases where knowledge of the actual therapy is vital for future patient care.

#### Trial sites

Division of Dermatology, Department of Medicine, King Chulalongkorn Memorial Hospital, Bangkok, Thailand.

#### Biological sample collection for genetic analysis

Biological sample collection includes 1 ml of saliva and a spoon of feces collected by participants in separate containers prepared by researchers and skin samples from three regions, two lesional skin samples, and one sample of nonlesional skin collected by skin taping by physicians. The five containers per individual containing nucleic acid preservation buffer will be transported to the laboratory in 30 minutes at room temperature or as soon as possible at 4 degrees Celsius. Upon arrival at the laboratory of the Faculty of Medicine, Chulalongkorn University, the sample will be extracted by Quick-DNA™ H M W MagBead Kit (cat. no. D6060, Zymo Research) and then amplified by the PCR method. The PCR amplification products will be purified by AMPure XP (Beckman Coulter) and the quantity will be measured by NanoPhotometer^®^ C40 (Implen, USA). After that, DNA will be combined with the solution for the Ligation Sequencing Kit (LSK-109, Oxford Nanopore Technologies) to perform DNA sequencing using the Nanopore MinION sequencing system. All samples will be recorded and barcoded with a unique storage ID.

#### Data collection and management

Data collection in this study is based on personal characteristics, clinical evaluations, and self-reported measures. Baseline demographic and clinical characteristics include age (years), sex, weight, body mass index (BMI), familial history of psoriasis, duration of psoriasis, comorbidities (diabetes mellitus, hypertension, dyslipidemia, obesity, metabolic syndrome, cardiovascular disease, psoriatic arthritis, inflammatory bowel disease, psychiatric disorders, among others) and concurrent therapies for systemic psoriasis. The local psoriasis severity index (LPSI), self-reported itching on the visual analog scale, reported adverse events, and microbial profile are collected at the time points below.

All investigators will have access to the final trial data set. All documents will be safely stored in the Skin Unit Research Facilities for Academic and Clinical Excellence (SURFACE), Division of Dermatology, Department of Medicine, Faculty of Medicine, Chulalongkorn University, for ten years. No identifiable data will be recorded and all documents will be recorded with the research identification number.

### Outcome measures

Evaluations are collected at four time points: 0^th^, 30^th^, 60th and 90^th^ day of the study. Fifty percent of the participants are chosen at random to have their saliva, feces, and skin samples collected if they consent to the storage and future testing of biological materials. An overview of all the assessment points and results is shown in **Table 2**.

**Table 2.**
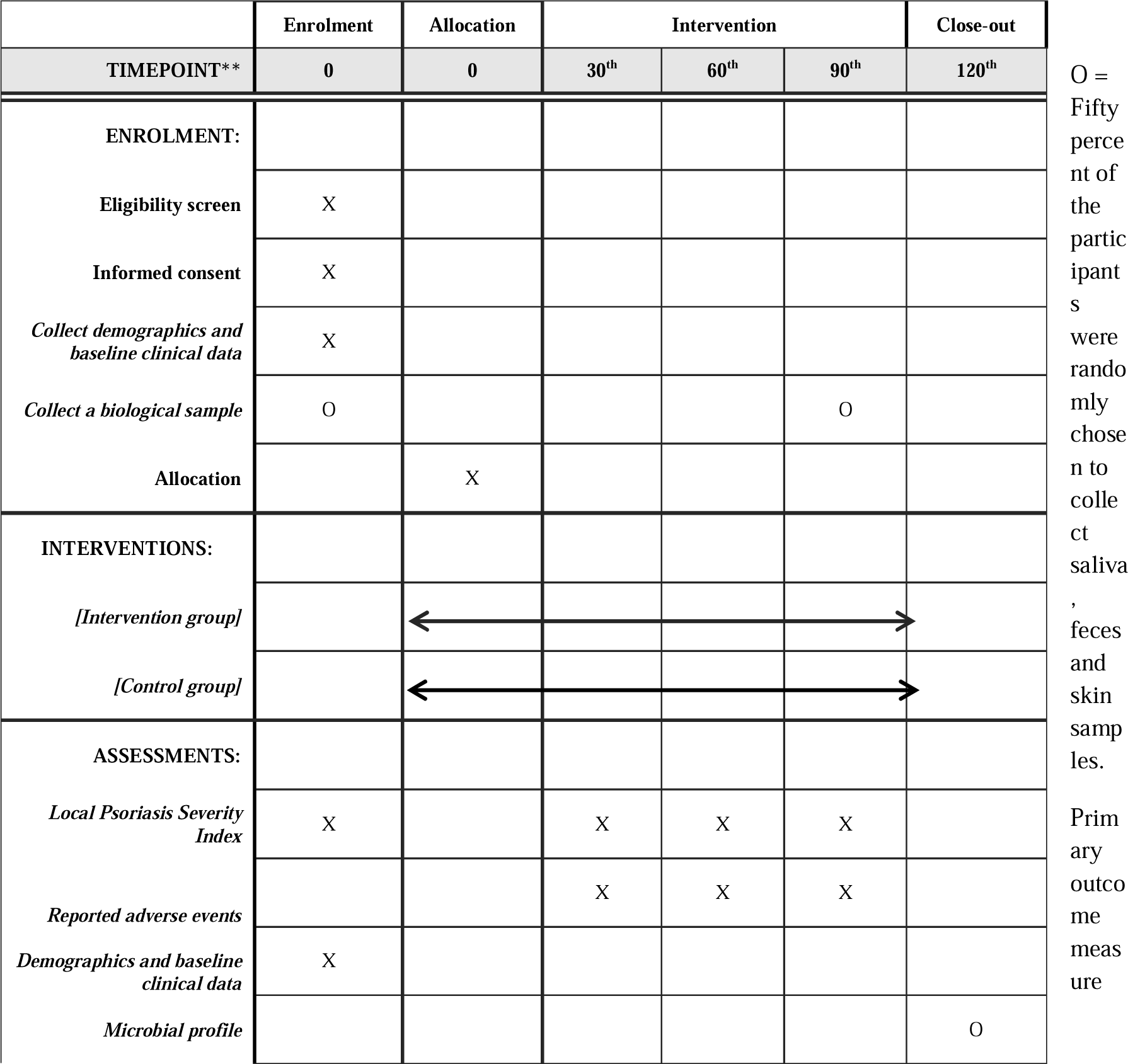
SPIRIT Schedule for enrollment, interventions, and assessments.

#### Local psoriasis severity index (LPSI)

Each lesion will be assessed with a modified psoriasis area severity index called the local psoriasis severity index (LPSI). The LPSI is the sum of the following symptoms evaluated by the physician: erythema (redness), induration (thickness) and desquamation (scaling infiltration). Each score was classified as follows: 0 = no symptoms, 1 = slight symptoms, 2 = moderate symptoms, 3 = marked symptoms, 4 = very marked symptoms [78]. P.A., a dermatology professor, conducts all LPSI evaluations for this study.

### Secondary outcome measure

#### The itch score by the visual analog scale

The visual analog scale consists of a line 10 cm long, with verbal anchors at either end. The patient places a mark on the line corresponding to the patient’s intensity rating. VAS is one of the most commonly used methods for assessing the severity of pruritus, as it provides an easy, rapid and valid estimate of the itch [79-81]. Participants will be asked to indicate the point within three minutes of this study.

#### The alpha-diversity, beta-diversity, and relative abundance of microbiota

Biological samples, including saliva, feces, and skin, will be analyzed for microbial alteration in the form of microbial diversity. The diversity of microbes will be defined as the proportion and abundance distribution of different types of organism. The abundance distribution includes alpha diversity, which is an abundance of different bacterial taxa in a single sample, and beta diversity, which is microbial diversity in different samples. Relative abundance is the ratio of the absolute abundance of a taxon to the total absolute abundance of all taxa in a unit volume of an ecosystem. [8, 82, 83]

### Reporting of adverse events

Participants are encouraged to contact research staff if they have concerns about mental or physical health decline. Upon the occurrence of an adverse event, the patient would receive treatment for the adverse event at King Chulalongkorn Memorial Hospital for free.

### Discontinuation and withdrawal

Patients who experience major adverse effects after enrolling in the study, participate less than 80% of the research period, or are unwilling to continue participating in the trial will be excluded. However, the data already collected and the reason for the cessation of study participation may be included in the final report.

## STATISTICAL ANALYSIS

### Sample size calculation

This study compares the mean LPSI score of transdermal CBD and placebo use. With the 80% power, α = 0.05, effect size 0.58, and allocation ratio 1:1, the determined sample size per group is 48 individuals. However, there is a potential for around 20% of the individuals to drop out of the study. The researchers determine that the total sample size is 60 individuals.

### Statistical method

Statistical analysis will be performed using STATA version 15.0 (StataCorp. 2017. Stata Statistical Software: Release 15. College Station, TX: StataCorp LLC.). Categorical data will be expressed as a number and depending on which is more appropriate, continuous data will be reported as mean +/- standard deviation or median +/- interquartile range. Qualitative variables will be presented in a table and then analyzed by the Chi-square test or Fisher’s exact test. The paired T-test or the unpaired T-test will be applied for quantitative data with normal distribution. The Wilcoxon signed ranks test or Mann-Whitney U test will be used for the quantitative data, which is not present with normal distribution.

### Sequencing and Microbiome data analysis

The taxonomic classification will be assigned for the V3/V4 16S region. All taxonomic classifications will be implemented within QIIME2. Bacterial diversity is determined by alpha diversity and beta diversity, which will be calculated by the QIIME 2 pipeline. Alpha diversity will be analyzed using Shannon’s diversity and Simpson’s diversity. Beta diversity will be analyzed by Bray-Curtis dissimilarity.

## TRIAL STATUS AND TIMELINE

The research will be advertised from April 2023 to May 2023. Subsequently, subject recruitment will occur from May 2023 to June 2023. The duration of the intervention will be between July 2023 and September 2023. Data analysis will be conducted from September 2023 to December 2023. The presentation of the data and the preparation of the manuscript will be completed by January 2024.

## DISCUSSION

This study will be the first randomized controlled study to assess the efficacy and safety of CBD-containing transdermal patches with minimal THC in patients with mild to moderate plaque-type psoriasis. It will also be the first to investigate the correlation of the baseline and alteration of the microbial profile and the efficacy and safety of CBD with minimal THC patches, which can help with customized patches, in addition to age, sex, BMI, and genetic variables. If the intervention shows significant positive results, the promise of minimal THC transdermal CBD patches as an alternative topical therapy option for psoriasis sufferers will be emphasized, and additional studies on a larger scale may be conducted on this issue. Further research could examine (i) the efficacy and cost-effectiveness compared to the standard of care, (ii) barriers to implementation (e.g. social stigma, cost of administration, and legalization policies)

The strengths of this study include (i) a double-blind randomized design, (ii) a review of the aspects of the patients and physicians of the psoriasis symptoms, (iv) transdermal patches that could better control the amount of CBD than the previous form of transdermal CBD administration, and (v) investigation of personalized factors that include the microbial profile and demographic and clinical characteristics. However, this study also has several limitations including (i) recruiting and providing intervention at a single site and in an academic medical center setting that could limit generalizability and (ii) the subjective nature of the itch score, which includes individual variation. The study results will be released to the participating physicians, patients, and the general medical community. For reproducible research, we will transfer a collection of entirely anonymized data to a suitable data archive for sharing purposes. Any protocol amendment will be approved by the Institutional Review Board of Chulalongkorn University and the Thai Clinical Trials Registry prior to implementation.

## Supporting information

SPIRIT 2013 Checklist

## Data Availability

All data produced in the present study are available upon reasonable request to the authors.

## Authors’ contributions

PS, KP, PA, PP, SV, TO, and SC conceived of the study and initiated the study design. PS wrote original draft. PS, KP, PA, and PP review and edit the draft. KP and PA supervised. PP provided statistical expertise in clinical trial design and primary statistical analysis. All authors contributed to the refinement of the study protocol and approved the final manuscript.

## Acknowledgements

None.

## Supporting Information

Supplementary 1. SPIRIT 2013 Checklist: Recommended items to address in a clinical trial protocol and related documents.

